# Cardiovascular events associated with CDK4/6 inhibitors based on randomized controlled trials or cohort trials: a safety meta-analysis

**DOI:** 10.1101/2024.03.29.24305099

**Authors:** Chengrong Zhang, Guoshuang Shen, Shengmei Li, Fei Ma, Huihui Li, Yuyao Tang, YongXin Li, Zhoujuan Li, Zijun Zhu, Tianlei Qiu, Zhilin Liu, Yi Zhao, Shifeng Huang, Fuxing Zhao, Fanzhen Kong, Jiuda Zhao

**Affiliations:** Breast Disease Diagnosis and Treatment Center of Affiliated Hospital of Qinghai University & Affiliated Cancer Hospital of Qinghai University, Xining, 810000, China; State Key Laboratory of Molecular Oncology, National Cancer Center/National Clinical Research Center for Cancer/Cancer Hospital, Chinese Academy of Medical Sciences and Peking Union Medical College, Beijing, 100021, China; Department of Breast Medical Oncology, Shandong Cancer Hospital and Institute, Shandong First Medical University and Shandong Academy of Medical Sciences, Jinan, 250117,China

**Keywords:** CDK4/6 inhibitors, adverse events, cardiovascular events

## Abstract

**Background:** CDK4/6 inhibitors is highly valued, but the incidence of cardiovascular events (CVAEs) associated with CDK4/6 inhibitors is not clear.

**Methods:** Eligible CVAEs were extracted from the ClinicalTrials.gov registry. A systematic search of electronic databases (PubMed, Embase, Cochrane Library, and important meetings) until 3 September 2023 was conducted. A disproportionality analysis was performed from the first quarter (Q1) of 2013 to Q1 of 2023 using data from the FDA Adverse Event Reporting System database. Study heterogeneitywas assessed using the *I^2^* statistic. Using Peto OR and inverse variance methods to calculate the risk and incidence of CVAEs associated with CDK4/6 inhibitors.

**Findings:** 21 RCTs and cohort trials (n=24,331) were included. During the follow-up period of 8.4 to 34.0 months, CDK4/6 inhibitors significantly increased the risk of CVAEs (Peto OR, 1.64, 95% confidence interval, 1.23 - 2.21, *P* < 0.01). The rates of QT prolongation and deep vein thrombosis were 98.83 (89.6-100.1) and 6.41 (5.23-7.18) per 1000 patients, respectively. Moreover, we identified 11 CVAEs that were not reported in RCTs or cohort studies, acute coronary syndrome, atrial fibrillation, and mobile thrombophlebitis etc. were strongly correlated with CDK4/6 inhibitors. Furthermore, the risk of CVAEs varied depending on the specific CDK4/6 inhibitors used, its combination with different endocrine therapies, and the patient’s treatment stage.

**Interpretation:** CDK4/6 inhibitors increase the risk of CVAEs, some of which may lead to serious consequences, early recognition and management of CVAEs is of great importance in clinical practice.

## Introduction

Breast cancer (BC) is the most commonly diagnosed malignancy and the primary cause of cancer-related deaths among women worldwide^1^. Clinically, standardized diagnosis and treatment significantly enhance patient prognosis^2,3^. However, new problems have emerged due to prolonged survival. In recent years, a large study found that patients with BC are at a higher risk of mortality from cardiovascular adverse events (CVAEs) than from the tumor itself^4–6^. Clinicians are increasingly paying attention to this prominent problem of CVAEs caused by antitumour treatments and urgently need to address it.

Multiple factors can lead to a high incidence of CVAEs in patients with BC. Firstly, lifestyle choices like like smoking, excessive alcohol consumption, and insufficient physical activity are considerable contributors^7^. Secondly, antitumour medications, such as endocrine drugs, immunotherapies, targeted therapies, and CDK4/6 inhibitors (CDK4/6 inhibitors) have been linked to CVAEs^8,9^. Furthermore, the risk of CVAEs, particularly with endocrine therapy, increases with prolonged treatment duration. Additionally, radiotherapy has been associated with CVAEs^10^. Thirdly, combining multiple cancer treatments can lead to increased heart toxicity^11^.

CDK4/6 inhibitors are esteemed for their efficacy in both early and advanced BC, particularly in large populations of patients with hormone receptor-positive (HR+) BC. In advanced stages, CDK4/6 inhibitors enhance progression-free survival (PFS) and overall survival (OS) in patients^12,13^. In early stages, studies such as MonarchE and NATALEE studies have indicated that CDK4/6 inhibitors significantly boost invasive disease-free survival (IDFS)^14,15^.

Several authoritative guidelines strongly recommend combining CDK4/6 inhibitors with endocrine therapy (ET) as either first- or second-line treatments for patients with HR+/HER2-advanced BC. Particularly, Abemaciclib is the standard recommendation for high-risk groups in early stages of the disease^16^. With the availability of CDK4/6 inhibitors, there has been a significant increase in their usage among HR+/HER2-BC patients^17^. The efficacy of CDK drugs in breast cancer treatment has not only led to a surge in the use of CDK4/6 inhibitors among these patients but also an extension in the duration of treatment. Concurrently, the associated toxicities of CDK4/6 inhibitors, the most important of which is cardiotoxicity, have gained significant attention. A recent randomized controlled trial (RCT) study highlighted that drug-induced cardiotoxicity can lead to premature morbidity and mortality^18^. Additionally, a real-world study utilizing data from the Florida Data Trust revealed that 24% of CVAEs occurred in the CDK4/6 inhibitors cohort, calculated at 35.9 per 100 person-years, marginally exceeding that in patients treated with anthracyclines^19^. Within the CDK4/6 inhibitors group, the mortality rate was notably higher for those with atrial fibrillation or heart failure^8^. However, no clinical trial system has been established to elucidate the incidence of CVAEs caused by CDK4/6 inhibitors. Evaluating cardiotoxicity is crucial for patients with cancer undergoing CDK4/6 inhibitors therapy, ensuring that cancer treatments do not compromise their long-term heart health^20^.

Therefore, we conducted a systematic review and meta-analysis of RCTs and prospective cohort studies to determine the risk of developing CVAEs associated with CDK4/6 inhibitors.

## Methods

### Literature Search Strategy

The study protocol was registered in advance with the International Prospective Register of Systematic Reviews (registration number: CRD42023462059). We used an integrated step-by-step approach to capture all available CVAE cases. Initially, we extracted all CVAEs categorized according to the Common Terminology Criteria for Adverse Events, Version 5.0 (CTCAE v5.0), from RCTs of CDK4/6 inhibitors listed on ClinicalTrials.gov. This extraction, conducted by two reviewers from inception until 30 July 2023 according to pre-specified selection criteria. Any discrepancies were resolved by consensus or negotiation with a third reviewer (JZ). Secondly, if ClinicalTrials.gov lacked CVAE reports, we then sourced data from published RCTs.

Concurrent electronic searches in PubMed and EMBASE utilized multiple keywords including cyclin-dependent kinase 4 and 6 inhibitors, palbociclib, ribociclib, abemaciclib, dalpiciclib, trilaciclib, cancer, neoplasms, breast cancer, breast tumors, cardiovascular toxicity, and cardiovascular adverse events. Search terms were applied to both subject headings and titles.Further searches included major oncology conferences of the European Society of Medical Oncology, the American Society of Clinical Oncology, the American Association for Cancer Research, the SAN Antonio Breast Cancer Symposium Database and ESMO Breast Cancer. Finally, in cases where CVAEs were unavailable on ClinicalTrials.gov or in published studie, we contacted the corresponding author or sponsor of the study via email to provide the requested information but did not receive a response.

We examined every identified RCT to avoid double counting, and only RCTs and cohort studies available for CVAES were retained in our final analysis. CVAEs of interest included deep vein thrombosis (DVT), hypertension, hypotension, QT prolongation, atrial fibrillation, myocardial infarction, cardiac failure congestive, cardiac failure, pericardial effusion, cardiac arrest, superior vena cava syndrome, embolism, phlebitis, tachyarrhythmia, ischaemia, supraventricular tachycardia, acute coronary syndrome, tachycardia, cardiomyopathy, palpitations, hypovolemic shock, angina pectoris, atrioventricular block, bradycardia, cardiopulmonary failure, ischaemic cardiomyopathy, and ventricular arrhythmia.

### Inclusion and Exclusion Criteria

The inclusion criteria were (1) phase 2 or 3 RCTs or cohort studies of HR-positive and HER2-negative BC, (2) randomised patients to receive CDK4/6 inhibitor plus ET or ET alone, any CDK4/6 inhibitor regimen, including a combination of different ET, patients at any stage, and (3) study of cardiovascular toxicity. The exclusion criteria were (1) phase 1 trials, (2) retrospective studies, (3) studies without cardiovascular toxicity, and (4) repetitive studies and reviews.

### Quality assessment

The quality of each included study was assessed using the Cochrane Collaboration’s ROB 2.0 tool to assess the risk of bias. Each study was evaluated for random sequence generation, allocation concealment, outcome assessment blinding, incomplete outcome data, selective outcome reporting, and other potential sources of bias. Each domain was assigned a ‘high’, ‘low’, or ‘unclear’ risk of bias independently by two reviewers, with disagreements adjudicated by a third reviewer. Sensitivity analysis was performed for all included studies. Funnel plots were used to assess publication bias.

### Study Objectives

The primary objective of our meta-analysis was to evaluate the the summary risk of CVAEs associated with CDK4/6 inhibitors exposure compared to control treatments. The treatment options for the control group included placebo, ET (including aromatase inhibitors [AI] or fluvestrant), placebo combined with ET, or chemotherapy of the physician’s choice.

The secondary objective focused on assessing the overall incidence of CVAEs. Three subgroup analyses were performed for different CDK4/6 inhibitors, CD4/6 Is combined with different ETs, and early stage and late-stage patients.

### Statistical analysis

A random-effects meta-analysis was performed to calculate the Peto Odds Ratio (OR) with a 95% confidence interval (CI). The incidence of CVAE was determined through a logit transformation and inverse variance weighting approach. Heterogeneity between studies was assessed using the *I*^2^ statistic of the inconsistency index and its *P*-value. According to the Cochrane Handbook of Systematic Reviews of Interventions, an *I*^2^ value >50% indicates substantial inter-study heterogeneity. Data management and meta-analysis of aggregate data (PETO method) were performed using R (4.3.1) and the R package Meta and are presented as forest plots. In all analyses, bilateral *P* < 0.05 was considered to indicate a statistically significant publication bias.

### Pharmacovigilance study in the FDA Adverse Event Reporting System (FAERS) database

A retrospective, heterogeneous pharmacovigilance study was conducted from the first quarter of 2013 (Q1) to the first quarter of 2023 (Q1) using FAERS to determine the risk of CDK4/6 inhibitor-CAVE in a large population. This study focused on CDK4/6 inhibitor, specifically palbociclib, ribociclib, abemaciclib, and trilaciclib. Dalpiciclib, not approved by the U.S. FDA, was excluded from the study. The study categorized reports in FAERS by System Organ Class (SOC), focusing on cardiac and vascular diseases, and collated clinical characteristics of CDK4/6 inhibitors-CVAEs patients, including sex, age, report year, region, and reporter type. Incomplete or incorrect patient records were excluded.

If the proportion of CVAEs was greater in cases than in non-cases, a disproportionality signal was considered. Utilizing a case/non-case approach akin to a case-control study, the reported odds ratio (ROR) method evaluated adverse reaction signals. If the number of CDK4/6 inhibitor-related adverse events exceeded 3 and the lower bound of the 95% CI for the ROR was greater than 1, this indicated a statistical association between CDK4/6 inhibitor and specific adverse events.

## Results

### Descriptions of included studies

The PRISMA flow diagram of the study selection is presented in Fig 1. Details of the study characteristics are presented in Table 1. We evaluated 6,845 records in the database, including studies, international conference reports, and reviews, and selected 245 potentially suitable articles for further abstract and title screening. Finally, we determined that 17 RCTs and 5 cohort studies published between 2016 and 2023 met the predefined criteria. A total of 24,331 patients from 22 studies were included, of which 24,147 were analysed for safety in the treatment groups with CDK4/6 inhibitors combined with ET (n = 12,934) and control groups with ET, placebo, or chemotherapy (n = 11,213).

**Fig 1.**
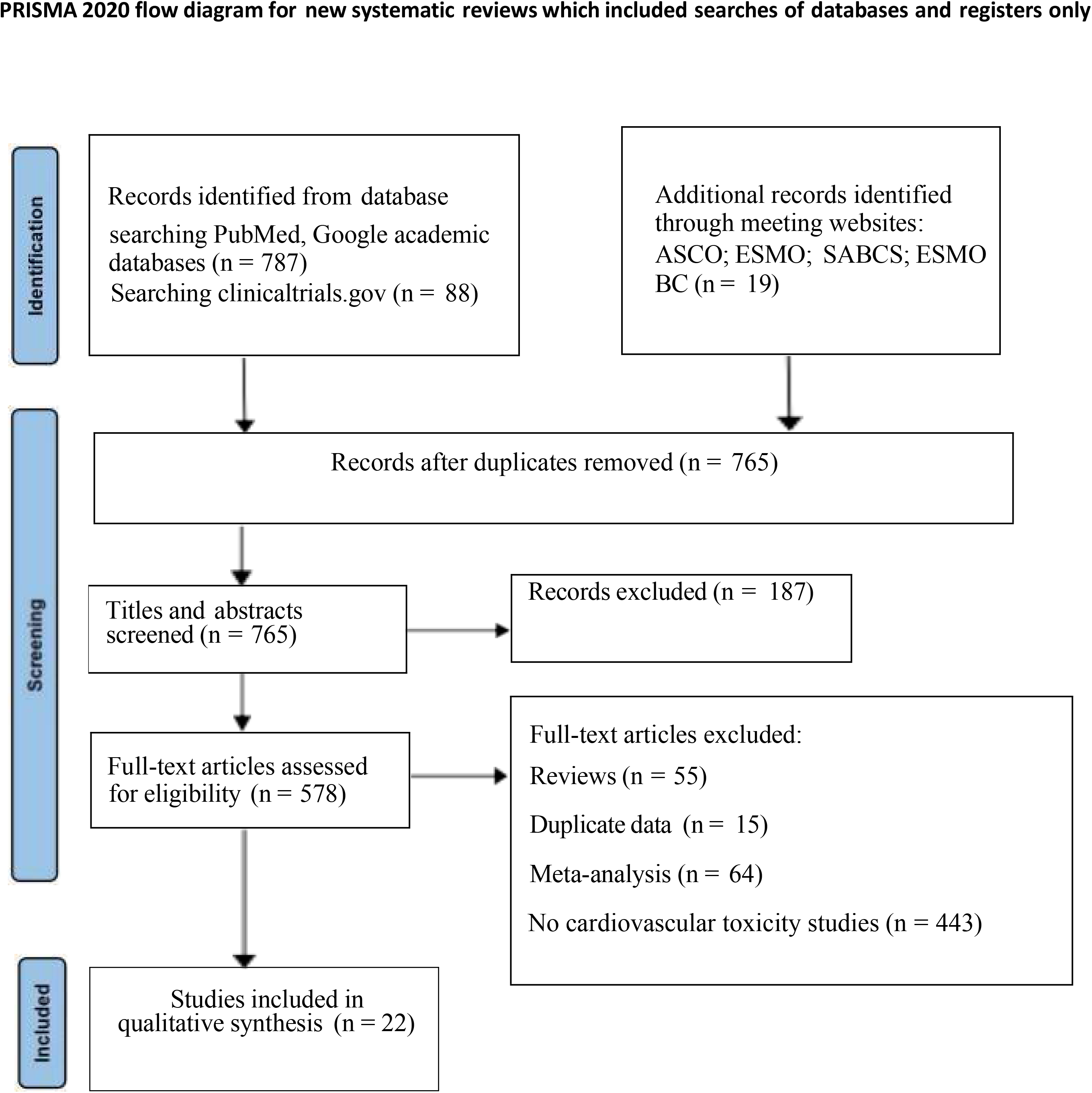
Search strings and flow charts for filtering and research selection.

**Table 1.**
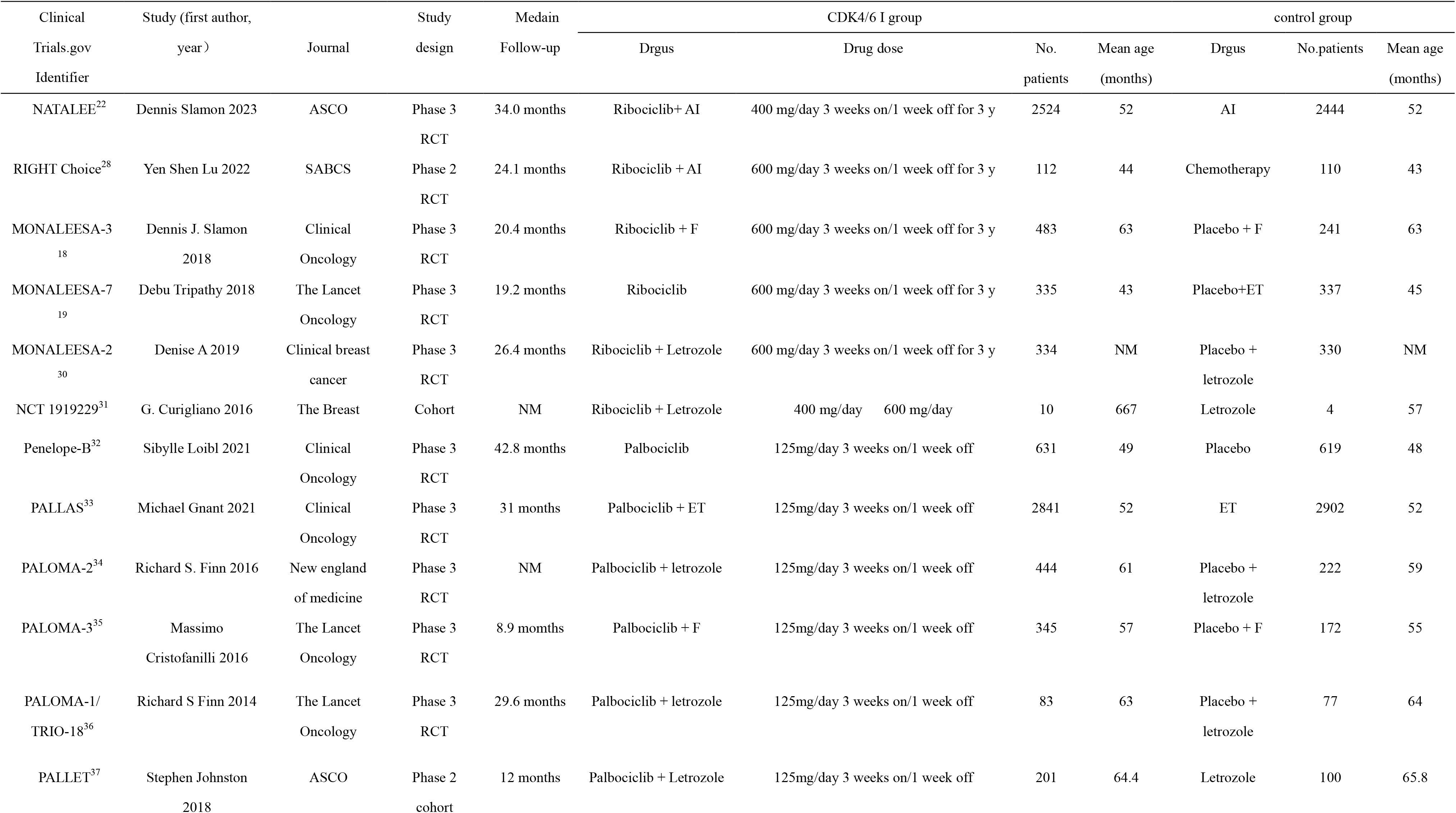
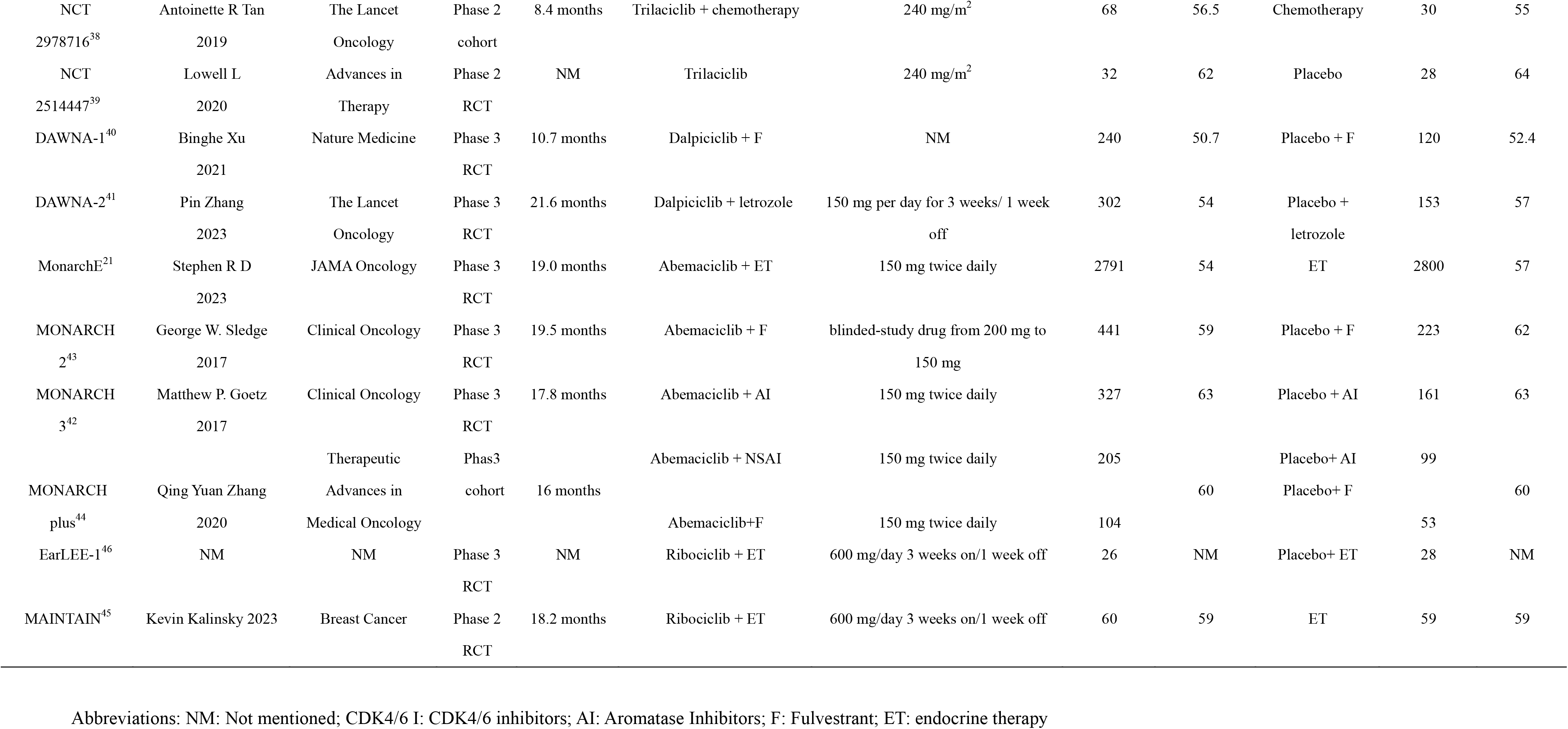
Characteristics of 22 trials included in this meta-analysis.

**Table 2.**
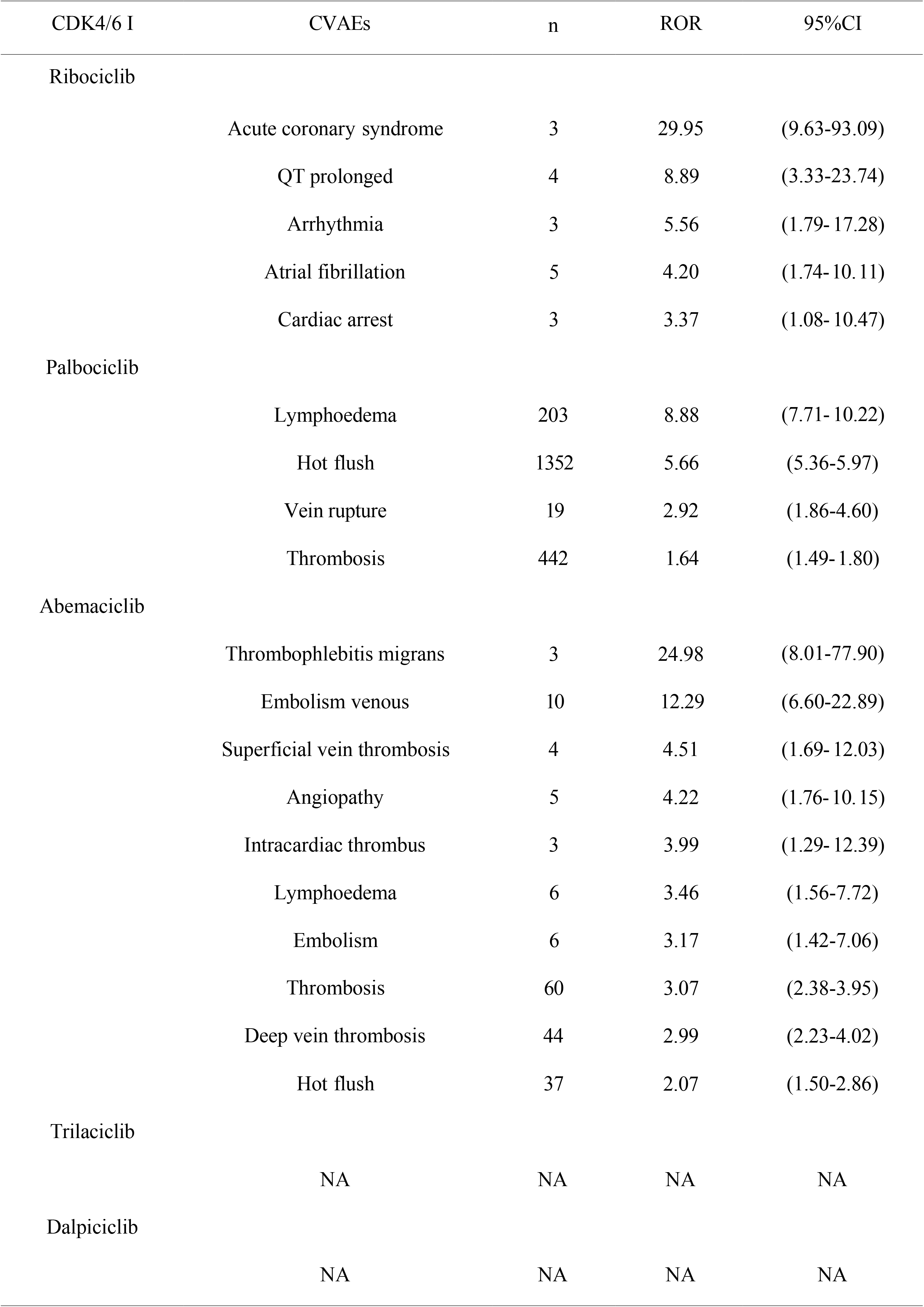
Disproportionality analysis in FAERS database.

| CDK4/6 I | CVAEs | n | ROR | 95%CI |
| --- | --- | --- | --- | --- |
| Ribociclib | Acute coronary syndrome | 3 | 29.95 | (9.63-93.09) |
|  | QT prolonged | 4 | 8.89 | (3.33-23.74) |
|  | Arrhythmia | 3 | 5.56 | (1.79- 17.28) |
|  | Atrial fibrillation | 5 | 4.20 | (1.74- 10.11) |
|  | Cardiac arrest | 3 | 3.37 | (1.08- 10.47) |
| Palbociclib | Lymphoedema | 203 | 8.88 | (7.71- 10.22) |
|  | Hot flush | 1352 | 5.66 | (5.36-5.97) |
|  | Vein rupture | 19 | 2.92 | (1.86-4.60) |
|  | Thrombosis | 442 | 1.64 | (1.49- 1.80) |
| Abemaciclib | Thrombophlebitis migrans | 3 | 24.98 | (8.01-77.90) |
|  | Embolism venous | 10 | 12.29 | (6.60-22.89) |
|  | Superficial vein thrombosis | 4 | 4.51 | (1.69- 12.03) |
|  | Angiopathy | 5 | 4.22 | (1.76- 10.15) |
|  | Intracardiac thrombus | 3 | 3.99 | (1.29- 12.39) |
|  | Lymphoedema | 6 | 3.46 | (1.56-7.72) |
|  | Embolism | 6 | 3.17 | (1.42-7.06) |
|  | Thrombosis | 60 | 3.07 | (2.38-3.95) |
|  | Deep vein thrombosis | 44 | 2.99 | (2.23-4.02) |
|  | Hot flush | 37 | 2.07 | (1.50-2.86) |
| Trilaciclib | NA | NA | NA | NA |
|  | NA | NA | NA | NA |
| Dalpiciclib | NA | NA | NA | NA |
|  | NA | NA | NA | NA |

### Risk of CVAEs associated with CDK4/6 inhibitor exposure

As shown in Fig 2A, CDK4/6 inhibitors significantly increased the risk of CVAEs compared with control treatment during a follow-up ranging from 8.4 to 34.0months (Peto OR, 1.66, 95% CI, 1.25 - 2.21, *P* < 0.01).

**Fig 2.**
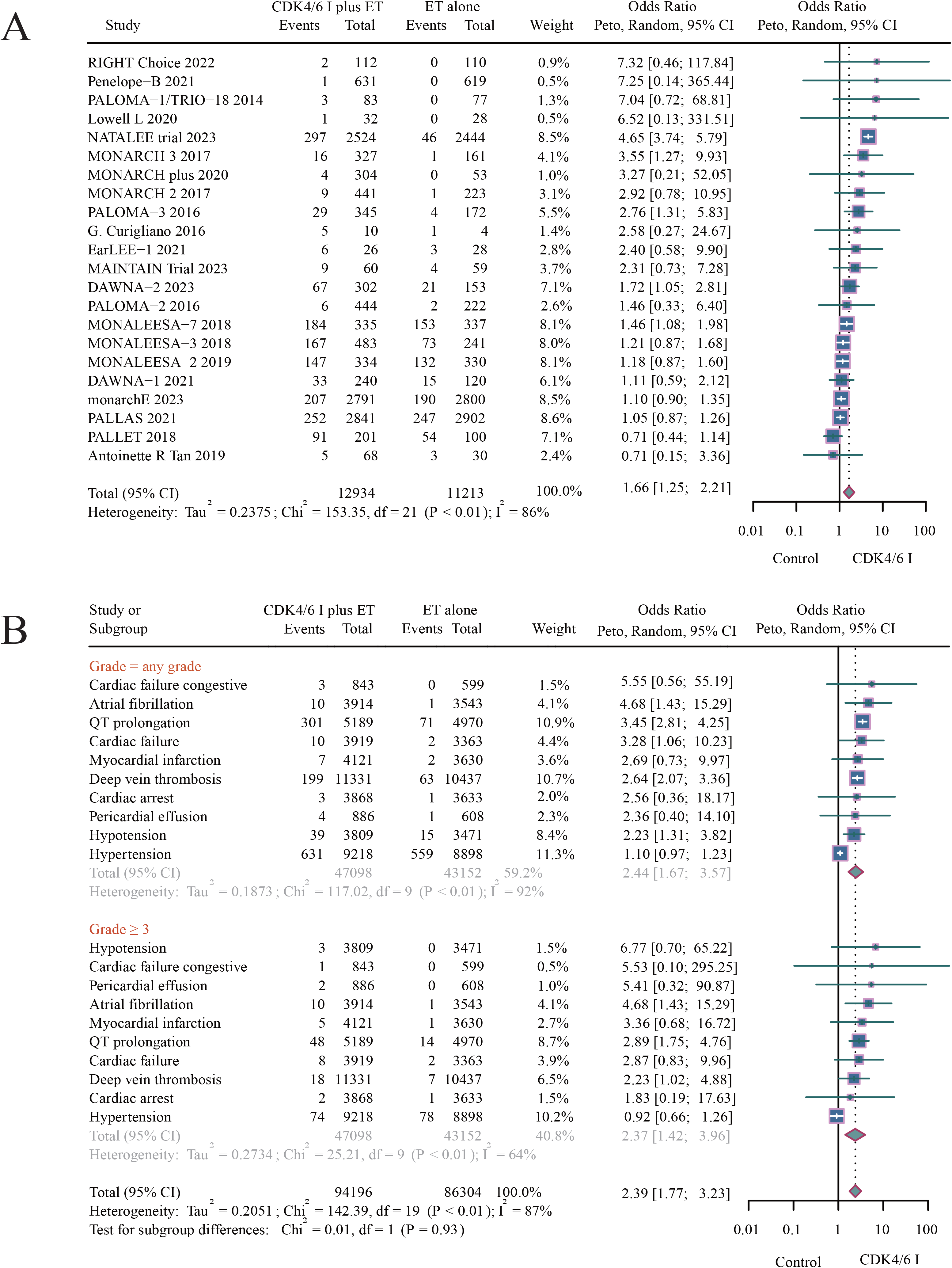
Peto OR of CDK4/6 inhibitors-CVAEs. (A) Summary pooled analysis forest plot on the Peto OR of CDK4/6 inhibitors therapy-associated CVAEs vs. controls. (B) Forest plot of analysis on type-specific CVAEs at any grade and grade ≥3 related to CDK4/6 inhibitors treatment vs. controls, Peto Odds Ratios.

As shown in Fig 2B, we included 28 different types of CVAEs, but only 10 CVAEs were derived from more than two studies. Therefore, we only calculated the Peto OR value of these 10 CVAEs. CDK4/6 inhibitors increased the risk of five CVAEs, including atrial fibrillation (Peto OR, 4.68, 95% CI, 1.43 to 15.29), QT prolongation (Peto OR, 3.45, 95% CI, 2.81 to 4.25), cardiac failure (Peto OR, 3.28, 95% CI, 1.06 to 10.23), DVT (Peto OR, 2.64, 95% CI, 2.07 to 3.36), hypotension (Peto OR, 2.23, 95% CI, 1.31 to 3.82).

### Incidence of grade ≥ 3 CDK4/6 inhibitor-related CVAEs

As shown in Fig 6A, statistical tests were conducted on adverse events of varying grades in a total of 18 RCTs or cohort studies. Compared to the control group, the combination of CDK4/6 inhibitors with ET is associated with a higher risk of CVAEs of grade ≥ 3 (Peto OR, 1.86, 95% confidence interval, 1.39 - 2.48, *P* < 0.01).

As shown in Fig 2B, CDK4/6 inhibitors increased the risk of three CVAEs of grade ≥ 3, including atrial fibrillation (Peto OR, 4.68, 95% CI, 1.43 to 15.29), QT prolongation (Peto OR, 2.89, 95% CI, 1.75 to 4.76), DVT (Peto OR, 2.23, 95% CI, 1.02 to 4.88).

### Incidence of CVAEs with an increased risk associated with CDK4/6 inhibitor **exposure**

27 different types of CVAEs were initially included in 22 RCTs and cohort studies, of which 17 CVAEs were from only two of the following studies, therefore, Peto OR were not calculated. Subsequently, this research quantified the incidence of 5 notable Peto ORs associated with CVAEs, including 58.01 (51.65-64.37) of QT prolongation per 1000 patients, 17.56 (15.14-19.98) DVT per 1000 patients, 10.24 (7.04-13.44) of hypotension per 1000 patients, 2.55 (0.97-4.14) of atrial fibrillation and cardiac failure per 1000 patients (Fig 3).

**Fig 3.**
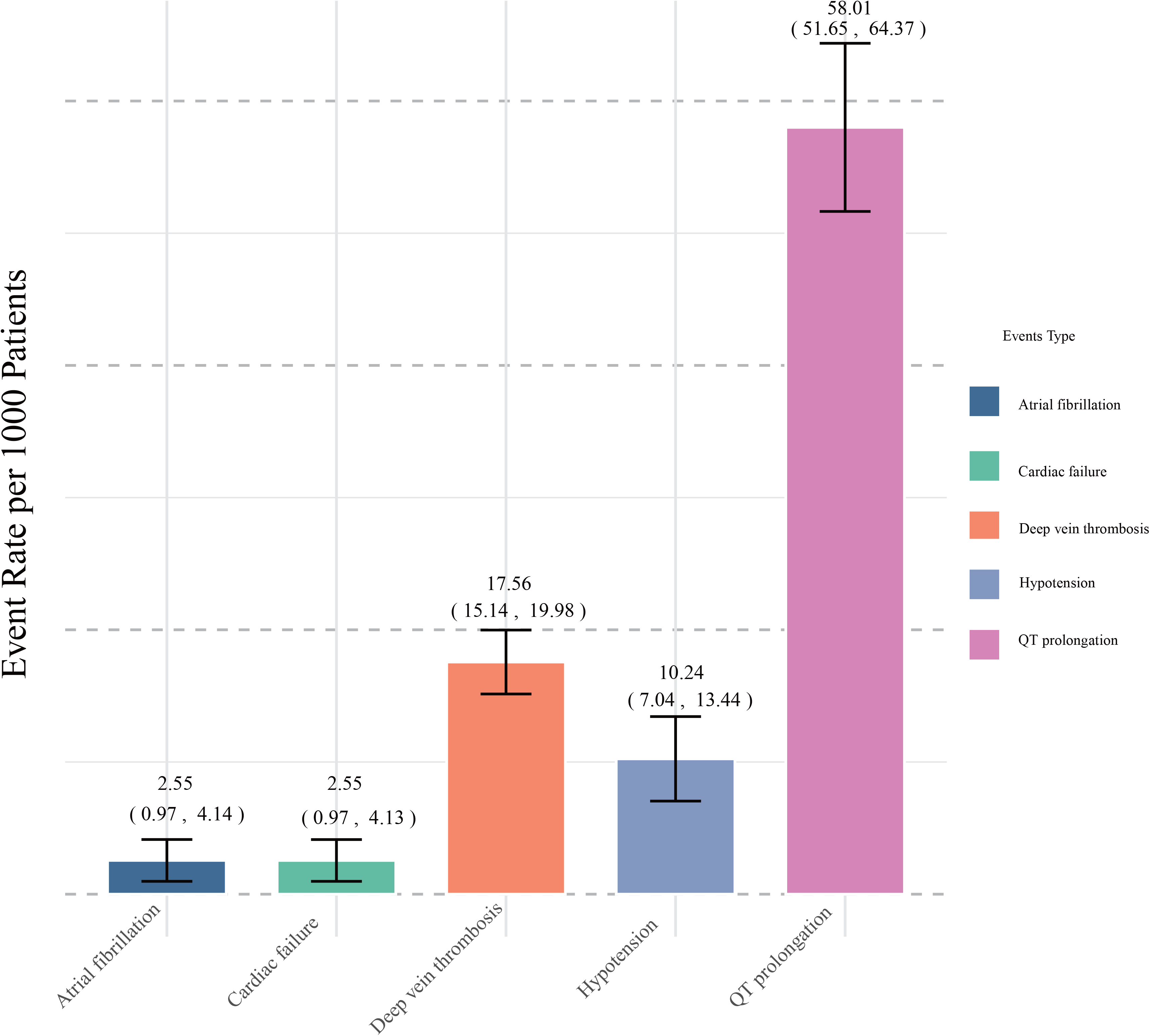
Summary pooled incidence analysis of CVAEs associated with CDK4/6 inhibitors therapy (per 1000 patients).

### Subgroups analysis

Among the five different CDK4/6 inhibitors, only ribociclib significantly increased the risk of CVAEs compared to the control group, while the remaining four CDK4/6 inhibitors did not show a statistically significant effect (Fig 4). For patients in different stages, use of CDK4/6 inhibitors in advanced stages increased the risk of developing CVAEs (Peto OR, 1.62, 95% CI, 1.28 to 2.06), while no statistical difference was seen in patients in early stages (Fig 5A). As shown in Fig 5B, compared to AI, the combination of CDK4/6 inhibitors with fulvestrant results in a higher risk of CVAEs (Peto OR, 1.94, 95% CI, 1.36 to 2.76). Figure 6B demonstrates that advanced patients with AI are more likely to develop CVAEs, while early patients are not statistically significant (Peto OR, 1.65, 95% CI, 1.20 to 2.28).

**Fig 4.**
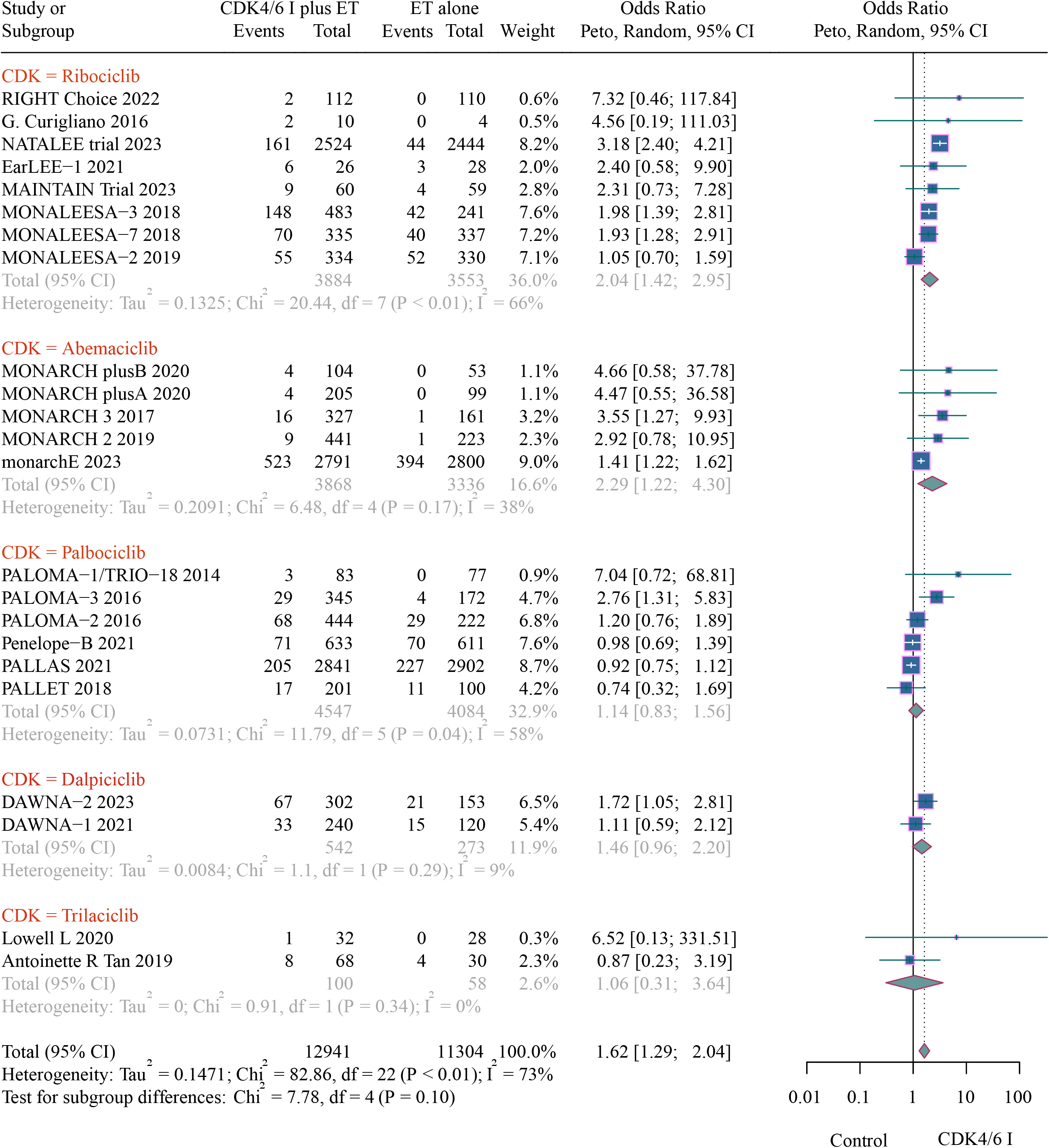
Subgroup analyses of association between CDK4/6 inhibitors and CVAEs according to different CDK4/6 inhibitors.

**Fig 5.**
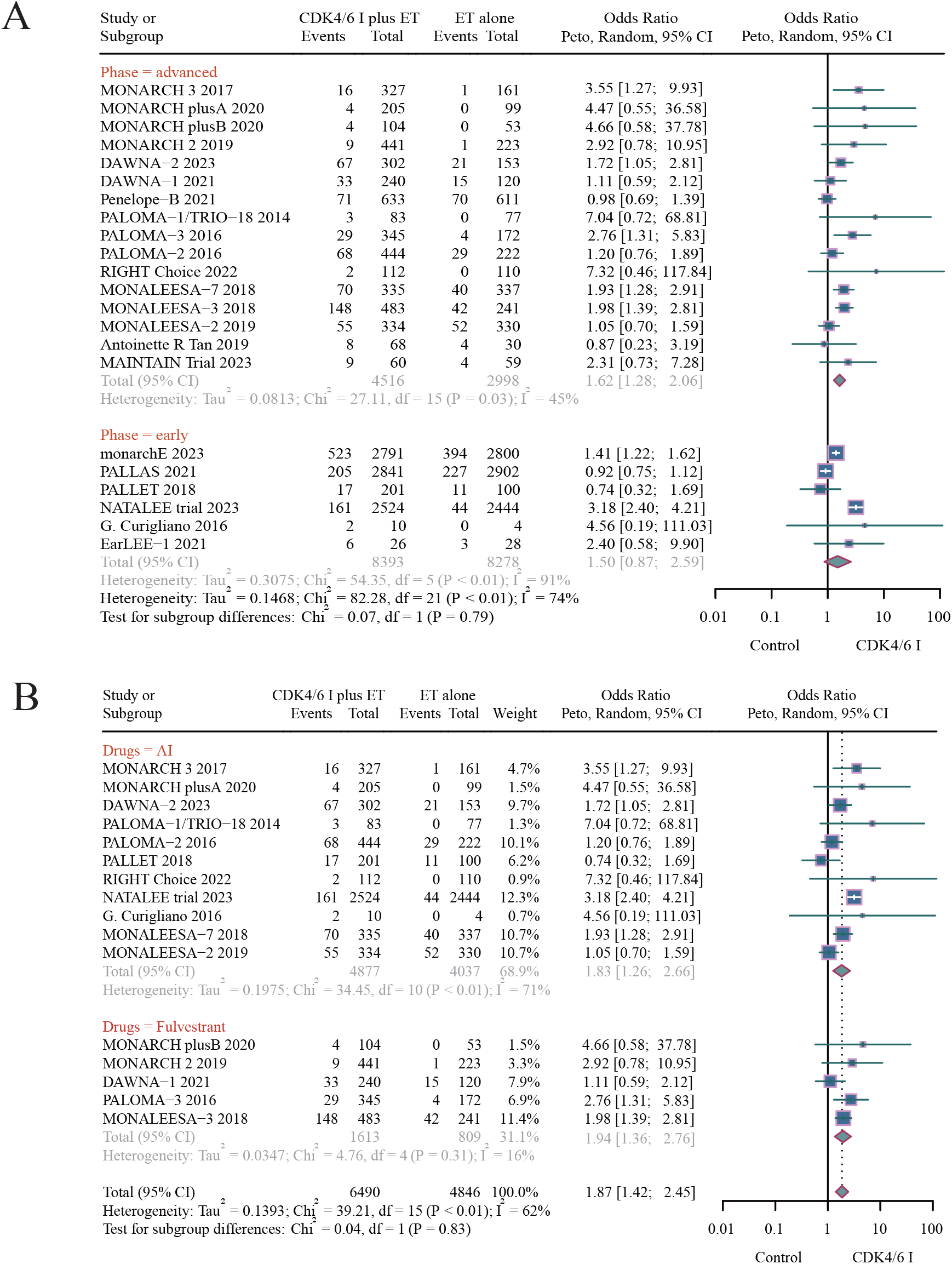
Subgroup analyses of association between CDK4/6 inhibitors and CVAEs. (A) According to the patients in different stages. (B) According to the different endocrine drugs combined with CDK4/6 inhibitors.

**Fig 6.**
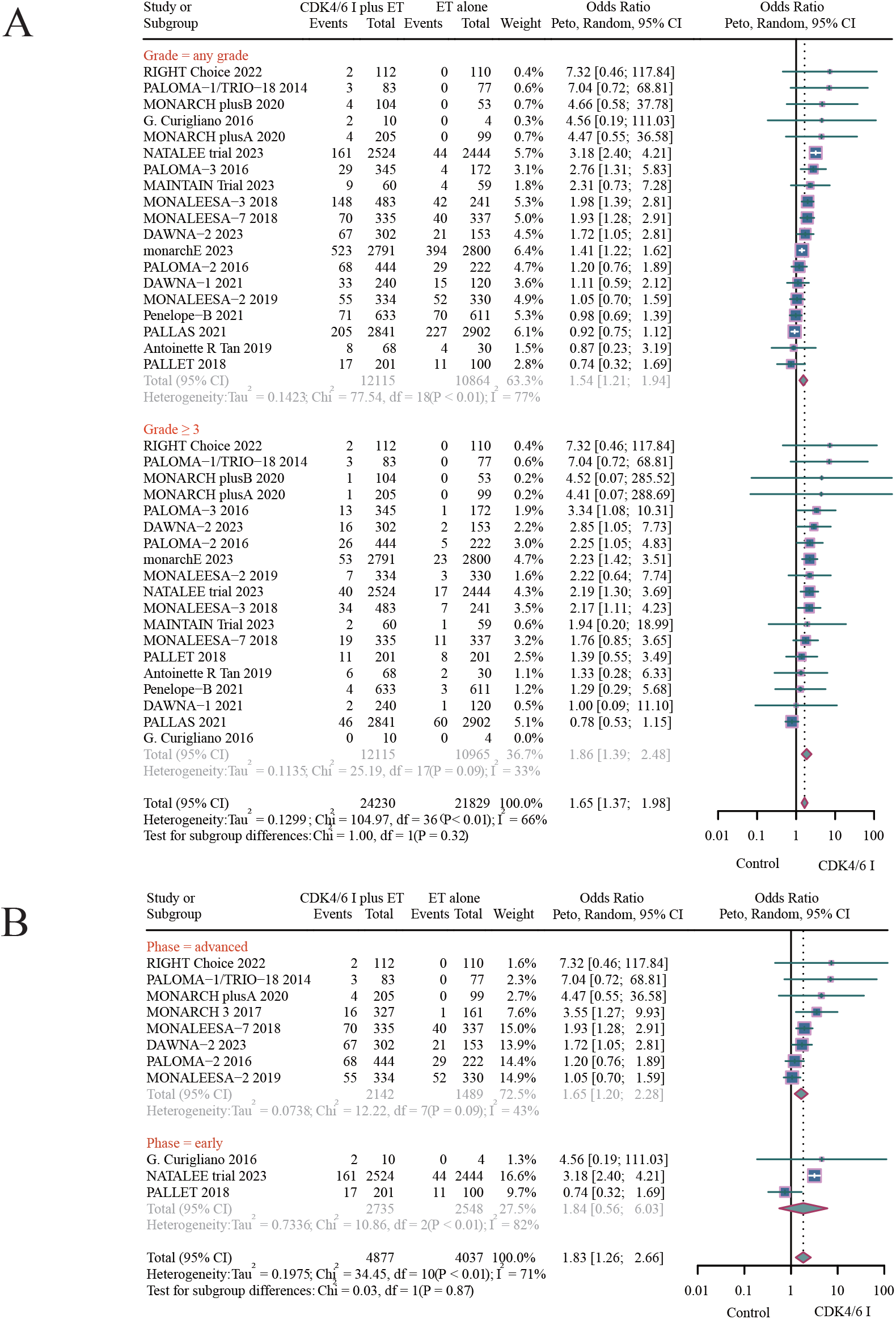
(A) Forest plot of analysis on the Peto OR at any grade and grade ≥3 related to CDK4/6 inhibitor treatment vs. controls. (B) Forest Plot of CVAEs in early and advanced breast cancer: CDK4/6 inhibitors and Aromatase Inhibitors vs. Control, Peto ORs.

### Characteristics of CDK4/6 inhibitor-related CVAEs recorded in FAERS

In total, 2,212 cases of CDK4/6 inhibitor-related CVAEs were identified in the FAERS database, including 18 cases for ribociclib (0.8%), 2016 cases for palbociclib (91.1%), and 178 cases for abemaciclib (8.0%). The characteristics of the CDK4/6 inhibitor-related CVAEs are presented in detail in Fig S1. The number of CDK4/6 inhibitor-related CVAEs reports increased dramatically from 2017 to 2022 (238 to 649 cases). The largest number with CVAEs was reported for albociclib (n = 2,016), whereas the smallest number of CVAEs was reported for ribociclib (n = 18).

The ROR disproportionate results are shown in Tabel 2, with the strongest signal being ribociclib and acute coronary syndrome (n = 3; ROR 29.95; 95% CI 9.63 to 93.09). Ribociclib was associated with QT prolongation (n = 4; ROR 8.89; 95% CI 3.33 to 23.74), which is consistent with the results of our meta-analysis. Palbociclib was significantly associated with lymphedema (n = 203; ROR 8.88; 95% CI 7.71 to 10.22). Abemaciclib is associated with thrombus and embolisms, including thrombophlebitis migrans,venous embolism, superficial vein thrombosis, intra-cardiac thrombus, embolism, thrombosis, and DVT. Trilaciclib did not have enough cases to calculate the ROR.

### Assessment of bias

The analysis of Cochrane risk assessment tools to estimate the risk of bias showed that the evaluation was the low risk (Fig S4 and Fig S5). We examined the funnel plot of the peto OR by visual inspection (Fig S3), which showed no significant asymmetry. Moreover, the results of the sensitivity analysis suggest a negative result, as can be seen in the supplementary material (Fig S2).

## Discussion

To our knowledge, this is the first comprehensive large-scale analysis of RCTs and cohort studies that comprehensively captured the association of CVAEs with CDK4/6 inhibitor based on 22 studies (n=24,331) and the FAERS database demonstrating that CDK4/6 inhibitor increases the risk of CVAEs. We made two major discoveries. First, CDK4/6 inhibitor increases the risk of QT prolongation, DVT, hypotension, atrial fibrillation and cardiac failure, and these CVAEs occur in 2.55 - 58.01 out of every 1,000 patients in RCTs or cohort studies treated with CDK4/6 inhibitor over a follow-up period of 8.4 to 34.0 months. Second, our disproportionate analysis of FAERS data identified seven CVAEs not reported in RCTs or cohort studies, warranting further investigation.

In recent years, six meta-analyses have investigated the adverse effects effects of CDK4/6 inhibitors. Of these, three studies compared their toxicity and feasibility^21–23^, while the remaining three focused on hematological toxicity, interstitial lung disease, and venous thrombosis risks^24,25^. Notably, CVAEs related to CDK4/6 inhibitors, with the exception of DVT and QT prolongation, have not been systematically analyzed. While some studies noted QT prolongation, its association with CDK4/6 inhibitors remains inconclusive. One study reported an increased attributable risk (AR) for QT prolongation with ribociclib^26^, but lacked systematic analysis and AR is suboptimal for evaluating rare events. Another study on DVT risk, using relative risk (RR) and risk difference (RD), suffered from small sample size issues, whereas the Peto Odds Ratio (OR) method is deemed more precise for binary studies involving rare events ^66^. To enhance our study’s robustness, we integrated real-world and clinical data by augmenting our dataset with information from the FAERS database.

Considering the heightened risk of cardiovascular toxicity in cancer patients, specialized cardiology guidelines have been formulated to cater to their specific requirements. These guidelines provide more targeted treatment approaches, focusing on the protection of cardiovascular health in individuals affected by cancer^27,28^. In our meta-analysis, most of the attention was paid to QT prolongation and DVT. CDK4/6 inhibitors were significantly associated with QT prolongation among the CVAEs of interest, and the strength of the association between ribociclib and QT prolongation was found in the FAERS database. The QT prolongation induced by CDK4/6 inhibitors may be related to three kinds of mechanisms, including altering the expression of QT-related genes (KCNH2, SCN5A, SNTA1), influencing potassium and sodium channels, and blocking hERG-encoded potassium channels^29,30^.

The high risk of DVT associated with CDK4/6 inhibitors found in this study resulted in a black-box warning from the FDA. The incidence rate of DVT was low relatively low (17.56 per 1000), yet it was approximately three times higher in patients treated with CDK4/6 inhibitors compared to controls, corroborating the findings of Kyaw et al^31^. Analysis of the FAERS database revealed that abemaciclib notably increases the risk of thrombosis, including superficial, visceral, and rare-site thrombosis. Many researchers attribute the high coagulation state in cancer patients as a primary cause of thrombosis. And somatic tumour mutations in CDKN2, a physiological inhibitor of CDK4/6, may heighten the risk of cancer-associated DVT. Abnormalities in the CDK4/6 pathway, frequently observed in HR+/HER2-patients, could also contribute to the thrombosis mechanism ^32^.

The incidence of atrial fibrillation and heart failure was observed to be significantly elevated in the CDK4/6 inhibitor (CDK4/6 inhibitor) group compared to the control group, these findings align with the results reported by Michael G. Fradley et al^33^. Research indicates that CDK4/6 inhibitor may directly impact the cardiovascular system, involving alterations in potassium and sodium channel activity, increased vascular inflammation, left ventricular remodeling, and downregulation of the PI3/AKT pathway, the occurrence of all these factors may be related to the development of cardiovascular disease^34,35^. Some studies have also noted an increased risk of hypotension associated with CDK4/6 inhibitors, which is consistent with our meta-analysis^36,37^. While the precise mechanisms remain unclear, but it is speculated that CDK4/6 inhibitors could regulate vasodilation and contraction by inhibiting certain stages of the cell cycle, thereby influencing blood pressure^38,39^.Additionally, Patients using CDK4/6 inhibitors showed higher overall CVAE rates than the control group, regardless of grade severity. Notably, all-grade CVAEs induced by CDK4/6 inhibitors exhibited a higher risk compared to grade ≥ 3 CVAEs.

Surprisingly, we found significant findings from the FAERS database. Consistent findings linked CVAEs like acute coronary syndrome and arrhythmia with CDK4/6 inhibitors. Events such as cardiac arrest and thrombosis also showed varying degrees of association. Intriguingly, certain events, while not statistically significant in meta-analyses, exhibited warning signs in the FAERS data, suggesting potential risks. Although not significantly tied to increased acute coronary syndrome risk, CDK4/6 inhibitor, especially ribociclib, demonstrated a strong association in the FAERS database, as observed with harrhythmia and cardiac arrests. We have two hypotheses for this situation, firstly, the real-world data might include patients with pre-existing cardiovascular conditions, as there are no stringent exclusion criteria. Secondly, patients in real-world scenarios might have been undergoing multiple antitumor therapies before starting CDK4/6 inhibitors, potentially elevating the incidence of CVAEs in the FAERS database.

Subgroup analyses indicated varied CVAEs linked to different CDK4/6 inhibitors. Ribociclib was associated with a higher risk of acute coronary syndrome and prolonged QT interval, although based on only 3 or 4 cases, the reliability of the data needs further study. Abemaciclib was notably linked to DVT, superficial vein thrombosis, and visceral vein thrombosis. Although palbociclib poses a certain level of warning risk, its ROR value was not significantly high in the FAERS database. Nonetheless, the cardiovascular risks associated with its use should not be overlooked. The CVAEs risks of trilaciclib and dalpiciclib, both in clinical research and the FAERS database, were inconclusive and warrant further investigation in more clinical cases.

Overall, our meta-analysis revealed that only ribociclib was linked to an elevated risk of CVAEs, while other CDK4/6 inhibitors were identified solely as cautionary indicators in the FAERS database. This outcome was closely related to the characteristics of the patient population under study. The FAERS database encompasses a wide range of individuals and provides large samples. These patients often present with complex medical conditions, including concurrent organ diseases; particularly, patients with cardiovascular and cerebrovascular diseases are more sensitive to adverse reactions to CDK4/6 inhibitor^40^. In contrast, RCTs and cohort studies were more likely to include patients with better baseline health and fewer comorbidities. For instance, RCTs such as the MONARCH, PALOMA, and MONALEESA series stipulated the inclusion of patients with normal organ function and rigorous exclusion criteria for those with visceral crises^16,41–43^.

In our meta-analysis, we discovered that patients treated with CDK4/6 inhibitor combined with fulvestrant had a higher risk of developing CVAEs than patients treated with AI, which increased risk was mainly due to the fact that patients treated with fulvestrant are mainly patients with advanced breast cancer, who are more prone to complications like CVAEs. AI treatment encompasses both early and advanced cases, with advanced patients showing a higher likelihood of CVAEs. However, the risk of CVAEs in early-stage patients receiving AI treatment is not statistically significant. Tamoxifen has been linked to an elevated risk of vascular events, such as thrombosis and lipid abnormalities^44^. while potentially reducing the risk of acute myocardial infarction and ischemic stroke ^45^. The concurrent use of tamoxifen with CDK4/6 inhibitors may influence the incidence of CVAEs. In the monarchE study control group, which included endocrine therapy with tamoxifen, the absence of specific data may have impacted the results^14^.

Patients in advanced stages of disease were observed to have a higher likelihood of developing CVAEs compared to those in early stages, a phenomenon potentially linked to the complex treatment regimens of advanced patients. Participants in studies like PALOMA-3, MONALEESA, and the DAWNA series were permitted to undergo targeted neoadjuvant therapy and endocrine therapy (ET), with some having previously received immunotherapy^13,43,46–48^. Simultaneously, targeted drugs (such as trastuzumab), ET drugs (such as letrozole and tamoxifen), and immune drugs (such as pabolizumab) may also cause contribute to the cardiovascular system damage, thereby increasing the risk of CVAEs^11,49^. Therefore, the heightened risk of CVAEs in patients with advanced disease can be attributed to both the complexity of their treatment and the progression of their disease.

### Strengths and limitations of the study

Our study had several strengths. Firstly, this review was pre-registered, prospectively registered, and reported in line with Preferred Reporting Items for Systematic Reviews and Meta-Analyses (PRISMA) 2. Secondly, we performed repeated screening and data extraction to assess the study-level risk of bias using the Cochrane risk of bias tool, risk of bias 2 (ROB 2), which is superior to ROB 1 in both rigor and objectivity^50^. Lastly, our data sources were extensive, and we specifically explored the association between CDK4/6 inhibitor and CVAEs using a dual approach of meta-analysis and pharmacovigilance disproportionality analysis, which is more persuasive when combining clinical trials with real-world data. The FAERS database is the FDA’s largest adverse drug reaction reporting system and this database is particularly suitable for the analysis of rare drug-related adverse events. By leveraging the wealth of information in the FAERS database, clinicians can better evaluate the possible associations between CDK4/6 inhibitor and adverse CVAEs.

However, this study has some limitations. Firstly, there was notable heterogeneity among the included studies, with the primary weaknesses being the variance in study design, follow-up duration, and types of CVAEs considered. Secondly, patients with different medical conditions have different degrees of CVAEs caused by different drug doses, which may affect the incidence of cardiovascular events. Thirdly, the FAERS database lacks detailed patient-level cancer characteristics, including hormone receptor status, prior chemotherapy exposure, endocrine therapy history, surgical or radiation history, and pre-existing cardiovascular conditions. The absence of these critical factors, crucial for determining disease severity, poses a challenge in accurately evaluating the incidence of CVAEs. Fourthly, some RCT studies only mention endocrine therapy based on standard control group criteria, without specifying the particular endocrine drugs. This lack of specific information prevents the subgroup analysis of different endocrine drugs in some studies, which may impact the assessment of the risk of CVAEs. Lastly, because of the large number of patients without a valid prescription, the FAERS database cannot accurately calculate the incidence of adverse events.

## Conclusion

CDK4/6 inhibitors are associated with an increased risk of QT prolongation, DVT, hypotension, atrial fibrillation and cardiac failure. Furthermore, the FAERS database identified seven additional CVAEs potentially linked to CDK4/6 inhibitors use, warranting further investigation. Risks vary with different inhibitors, combinations with ET, and patient treatment stages. These findings highlight the critical importance of enhanced cardiovascular monitoring for patients with breast cancer undergoing CDK4/6 inhibitors therapy.

## Contributors

Chengrong Zhang, Shengmei Li contributed to search studies. Drs Chengrong Zhang, Fanzhen Kong, Huihui Li, Yuyao Tang, YongXin Li contributed to extract data. Drs Chengrong Zhang, Guoshuang Shen, Jiuda Zhao contributed to finish the draft. All authors were involved in the revision and approval of the manuscript. Concept and design: Drs Chengrong Zhang, YongXin Li, Fei Ma, Jiuda Zhao. Acquisition, analysis, or interpretation of data: Zhoujuan Li, Zijun Zhu, Tianlei Qiu, Zhilin Liu. Drafting of the manuscript: Chengrong Zhang, Jiuda Zhao. Critical revision of the manuscript for important intellectual content: All authors. Statistical analysis: Yi Zhao, Shifeng Huang, Fuxing Zhao. Administrative, technical, or material support: Shengmei Li, Fei Ma, Fuxing Zhao, Yi Zhao. Supervision:Drs Jiuda Zhao. The corresponding author attests that all listed authors meet authorship criteria and that no others meeting the criteria have been omitted.

## Data Availability

YES

